# Gene regulatory programs of cognitive resilience and pathogenesis in Alzheimer’s disease

**DOI:** 10.64898/2026.02.19.26346666

**Authors:** Collin Spencer, N.M. Prashant, PsychAD Consortium, Jaroslav Bendl, Gabriel E. Hoffman, John F. Fullard, Donghoon Lee, Panos Roussos

**Affiliations:** Center for Disease Neurogenomics, Icahn School of Medicine at Mount Sinai, New York, NY, USA; Department of Psychiatry, Icahn School of Medicine at Mount Sinai, New York, NY, USA; Department of Genetics and Genomic Sciences, Icahn School of Medicine at Mount Sinai, New York, NY, USA; Friedman Brain Institute, Icahn School of Medicine at Mount Sinai, New York, NY, USA; Mental Illness Research, Education and Clinical Center VISN2, James J. Peters VA Medical Center, Bronx, NY, USA; Center for Precision Medicine and Translational Therapeutics, James J. Peters VA Medical Center, Bronx, NY, USA

**Keywords:** Alzheimer’s disease, cognitive resilience, gene regulatory networks, transcription factors, single-nucleus RNA-seq, NF-κB, BCL6, FLI1, IKZF1, IRF8, microglia, network rewiring, dorsolateral prefrontal cortex

## Abstract

**INTRODUCTION:** Cognitive resilience in Alzheimer’s disease (AD), wherein individuals maintain cognition despite substantial neuropathology, implies protective regulatory programs that remain poorly characterized.

**METHODS:** We constructed a cell-type-resolved gene regulatory network atlas from 1.7 million nuclei across 687 individuals spanning 26 cell types in the dorsolateral prefrontal cortex, classified as Control, Resilient, or AD dementia.

**RESULTS:** Analysis of 223 transcription factor regulons reveals a three-state regulatory framework: homeostatic erosion of IRF8/STAT1 interferon programs (State I), compensatory NF-κB suppression via BCL6 that distinguishes resilient from demented individuals (State II), and pathogenic FLI1/IKZF1 network expansion driving vascular-immune remodeling (State III). NF-κB emerges as the central hub, with BCL6-mediated repression and FLI1/RELA-driven activation constituting opposing molecular switches.

**DISCUSSION:** Replicated across independent cohorts, these findings model resilience as an active regulatory state and nominate stage-specific therapeutic strategies: restoring homeostatic programs, prolonging compensatory suppression, and constraining inflammatory escalation.

## 1. Introduction

Despite decades of research into the pathological hallmarks of Alzheimer’s disease (AD), Amyloid-β (Aβ) plaques and neurofibrillary tangles [1–3], the molecular mechanisms governing disease-associated differences and cognitive decline remain incompletely understood. A central paradox in AD research is the phenomenon of cognitive resilience [4–8], wherein individuals with substantial neuropathological burden maintain cognitive function, while others with similar pathology develop dementia. This discordance between pathology and clinical phenotype suggests that protective regulatory programs may buffer against neurodegeneration [6, 7].

While genome-wide association studies have identified AD risk loci [9–11] and single-cell transcriptomic analyses have cataloged cell-type-specific gene expression changes [12–15], these approaches largely fail to capture the higher-order regulatory logic – the coordinated activity of transcription factors (TFs) and their target genes – that orchestrates cellular responses to pathology. Prior efforts to infer gene regulatory networks (GRNs) in AD have been constrained by limited sample sizes, focus on individual cell types, or inability to compare disease, resilient, and control states within a unified framework [16, 17].

To address these limitations, we constructed one of the largest AD and cognitive resilience GRN studies to date, harmonizing regulatory inference across demographically diverse cohorts. By systematically comparing network architecture across AD, Resilient, and Control individuals using large-scale single-nucleus RNA sequencing (snRNA-seq) from the dorsolateral prefrontal cortex (DLPFC), we identify resilience-activated TF programs, quantify network rewiring dynamics, and characterize transcriptional differences across group contrasts. Our analysis reveals a three-state framework of transcriptional dysregulation in AD: State I (Homeostatic Erosion), State II (Resilient Compensation), and State III (Pathogenic Escalation). These designations reflect distinct regulatory profiles observed cross-sectionally rather than confirmed chronological stages of disease progression. This framework positions cognitive resilience as a dynamic regulatory state with therapeutic intervention potential, with BCL6 acting as a molecular switch whose activity distinguishes resilience from pathogenesis.

## 2. Methods

### 2.1. Human biosamples, ethics, and metadata harmonization

All brain specimens were obtained through informed consent via brain donation programs. This study was approved by the Institutional Review Boards of the Icahn School of Medicine at Mount Sinai, the National Institute of Mental Health, and Rush University Medical Center. The PsychAD cohort comprises 1,494 donors from three brain banks: the Mount Sinai NIH Neurobiobank (MSSM; 1,042 samples), the NIMH Intramural Research Program Human Brain Collection Core (HBCC; 300 samples), and Rush Alzheimer’s Disease Center (RADC; 152 samples) [18–20]. The cohort spans diverse genetic ancestries [21], ages, sexes, and neuropathological stages, enabling analysis across the full clinical spectrum from cognitively normal to severe dementia. Clinical and pathological metadata, including CERAD neuritic plaque density [22], Braak staging, and cognitive status, were harmonized across sources (see *2.2.1. Measuring cognitive impairment* for scale definitions and group criteria). CERAD scores were recoded onto an ordinal scale in which higher values correspond to greater neuritic plaque burden: 1 = no neuritic plaques (normal), 2 = sparse (possible AD), 3 = moderate (probable AD), and 4 = frequent (definite AD). Demographic and technical covariates (age, sex, *APOE* genotype, postmortem interval, batch) were collected per cohort.

### 2.2. PsychAD GRN cohort definition and clinical classification

From the PsychAD dataset spanning the MSSM and RADC cohorts, we defined mutually exclusive AD, Resilient, and Control phenotypes based on harmonized clinical and neuropathological metadata. To isolate canonical AD-associated regulatory mechanisms, individuals with co-occurring psychiatric, neurodevelopmental, or non-AD neurodegenerative diagnoses (e.g., diffuse Lewy body disease, frontotemporal dementia, or vascular dementia) were excluded.

A total of 250 individuals met criteria for the AD group, characterized by substantial proteinopathy burden and cognitive impairment, defined by CERAD = 4, Braak stage 3-6, and a consensus clinical diagnosis of dementia. The Resilient group included 95 individuals with comparable neuropathological burden (CERAD ≥ 3, Braak stage ≥ 3) but preserved cognitive function (no clinical dementia), representing individuals with neuropathological AD who nonetheless retained cognition. Finally, the Control group comprised 342 individuals lacking both significant Aβ or tau pathology (CERAD = 1, Braak stage ≤ 2) and any clinical diagnosis of dementia. Analyses were restricted to donors aged ≥65 years at death; within this age range the three groups showed comparable distributions of age, sex, and post-mortem interval, with age and sex additionally included as covariates in all downstream regression models.

#### 2.2.1. Measuring cognitive impairment

For the AD, Resilient, and Control contrasts defined above, a harmonized three-level ordinal variable of cognitive status served as the primary clinical phenotype. For MSSM donors, cognitive status was based on the Clinical Dementia Rating (CDR) scale (0 = no dementia; 0.5 = questionable dementia (very mild); 1 = mild; 2 = moderate; 3 = severe; 4 = profound; 5 = terminal) [23, 24]. For RADC donors, cognitive status was based on the physician’s overall cognitive diagnostic category (cogdx), assigned by a neurologist with dementia expertise at time of death, blinded to postmortem data, with case-conference consensus applied to selected cases; cogdx is coded 1-6 (1 = no cognitive impairment [NCI]; 2 = mild cognitive impairment [MCI] with no other cause of cognitive impairment; 3 = MCI with another cause; 4 = Alzheimer’s dementia with no other cause (NINCDS probable AD); 5 = Alzheimer’s dementia with another cause (NINCDS possible AD); 6 = other primary dementia) [18–20]. These source scales were collapsed into three ordinal levels used throughout: 0 indicates no dementia (CDR = 0 or cogdx = 1), 0.5 indicates mild cognitive impairment (CDR = 0.5 or cogdx = 2 or 3), and 1.0 indicates definite clinical dementia (CDR ≥ 1 or cogdx = 4, 5, or 6).

From this cohort definition, we constructed a PsychAD GRN subset consisting of 687 individuals (AD + Resilient + Control) and 1.7 million high-quality nuclei from DLPFC tissue. Gene expression matrices were filtered to the top 6,000 highly variable genes, as previously defined across the full PsychAD atlas following scanpy best practices for single-nucleus data normalization and variance stabilization. This subset formed the foundation for cell type-resolved GRN inference and downstream integrative analyses of pathogenic versus protective regulatory programs. Of the 27 subclasses in the PsychAD reference taxonomy, we excluded EN_L5_ET from all downstream analyses because it had fewer than 10,000 nuclei after preprocessing; the remaining 26 subclasses are used throughout this stud

### 2.3. PsychAD GRN inference and computational analysis

#### 2.3.1. Gene regulatory network inference

We inferred GRNs with the pySCENIC (v 0.12.1) pipeline [25, 26] across the PsychAD cohorts, including MSSM and RADC, and an external published dataset (Mathys et al. [4]) for each phenotype subset (AD, Resilient, and Control) independently using default parameters. We followed the standard SCENIC expression preprocessing; log-normalizing expression counts and selecting highly variable genes (3,192 total) while accounting for batch correction between datasets with scanpy (v1.9.3). pySCENIC’s GRNboost2 (arboreto v0.1.6) method was utilized for GRN inference. pySCENIC’s cisTarget function with Human motif databases v10 (https://resources.aertslab.org/cistarget/motif2tf/motifs-v10nr_clust-nr.hgnc-m0.001-o0.0.tbl) was used with default settings to prune regulon cis-gene targets using TF motifs within the promoter region of target genes. Due to the stochastic nature of GRNboost2, we performed GRN inference five times in each subset using randomized seeds and only retained consensus edges that appear in all five runs while averaging edge weights across all runs. Regulons, TFs and their target genes, were further pruned such that self-targeting regulons and those with fewer than 10 targets were removed. To ensure generalization of GRNs, we retained target genes that appear in MSSM, RADC, and an independent external cohort (Mathys et al. [4]) and averaged their edge weights, creating a finalized GRN for each phenotype (AD, Control, and Resilient).

#### 2.3.2. Regulon activity enrichment

We performed enrichment of regulons across cells using the AUCell python package and Regulon Specificity Score package with default parameters. To compare regulon enrichment between cell types, the resulting AUCell matrices were z-score normalized. We then used Robust Rank Aggregation (RRA), a statistical method that integrates multiple ranked lists to identify elements consistently ranked higher than expected by chance, applied to AUCell, RSS, and gene expression matrices to prioritize regulons that have differential enrichment across cell types. To assess concordance in regulon enrichment across cell types among MSSM, RADC, and the external Mathys et al. [4] cohorts, we computed the Pearson correlations between normalized z-scores using the pandas corrwith function. For each dataset, we computed the normalized regulon enrichment z-score and performed a meta-analysis between two datasets using Stouffer’s method.

#### 2.3.3. Differential regulon activity analysis

We applied Dreamlet for differential regulon activity analysis across different measures of AD pathology: case-control diagnosis, Braak, CERAD, dementia diagnosis, cognitive decline rating, cognitive and tau resilience, and cognitive and Aβ resilience. Building from the previously developed statistical tool Dream [27], dreamlet [28] applies linear mixed models to the differential expression problem in single-cell omics data. It starts by aggregating cells by the donor using a pseudobulk approach and fits a regression model to AUCell regulon activity. We aggregated AUCell non-count data using the function aggregateNonCountSignal. For each AD pathology measure, AUCell regulon activity, and cell cluster, the following mixed model was applied: AD Response Variable ∼ AD Clinical Variable + Sex + scale(Age) + log(N_genes) + Brain_bank + 1, where categorical and numerical variables were modeled as random and fixed effects, respectively.

To test whether there is a significant association between the target genes of a TF and disease signatures in a cell type, we performed Fisher’s exact tests between SCENIC GRN target genes and AD risk and Resilient gene signatures, based on Dreamlet differentially expressed gene (DEG) analysis. Disease genes with FDR < 0.05 were selected based on four different measures of AD severity, namely, case-control diagnosis, Braak, CERAD, and dementia status. The top 3 regulons were prioritized based on the overall enrichment of the AD DEG signature. The similarity between target genes of regulons was evaluated using the Jaccard similarity index.

#### 2.3.4. GRN rewiring analysis

To further prioritize regulons, TFs, and target genes involved in AD, we performed network analysis on the AD, Resilient and Control GRNs constructed in previous steps, calculating changes to the GRN structure based on the network architecture. We began by filtering for the top 25% of all edges in each phenotypic network (AD, Resilient, and Control), removing self loops, and filtering out TFs with fewer than 10 target genes to prioritize the most relevant TFs and target genes. Per TF, we subsequently calculated: the change in number of edges between GRNs, the overall change in importance (defined as the aggregated importance of the TF-gene edge weights), and changes to four measures of node importance (pagerank, degree centrality, betweenness centrality, and closeness centrality). For calculating centrality scores, we used the networkx v3.1 python package and their respective functions (pagerank, degree_centrality, betweenness_centrality, closeness_centrality) with standard parameters.

#### 2.3.5. Network visualization

Node centrality was calculated using PageRank analysis, which measures a ranking of the nodes in the graph based on the structure of the incoming links. The mean estimate and −log₁₀FDR of target risk genes in SCENIC regulons were visualized using the importance score edge weights from GRNboost2 with networkX (v3.1). For each regulon, SCENIC GRN TF target genes after cisTarget pruning were obtained and, using gseapy (v1.0.5), tested for gene-set enrichment based on the Gene Ontology Biological Processes 2023 [29, 30]. GO terms were clustered based on their Ward distance between −log_₁₀_FDR values; for the IRF1 differential regulon analysis **(Supplementary Figure 3)**, −log_₁₀_(P-value) was used in place of −log_₁₀_FDR and terms were required to overlap with ≥ 2 regulon genes.

#### 2.3.6. Cell-type GRNs

Cell-type GRNs for each major phenotype (AD, Resilient, and Control) were defined leveraging dreamlet differential regulon activity and AD-associated DEG results, as previously described. We pruned GRNs from each major phenotype based on regulon FDR < 0.05 significance across measures of AD in each given cell type, alongside target gene FDR < 0.05 significance from the Lee et al. [31] AD DEG analysis, resulting in cell-type-specific GRNs per phenotype. We conducted this stringent filtering along with preceding filtering steps to ensure high-confidence gene regulatory interactions, as false positives are a known limitation of GRN inference based on single-cell gene expression.

## 3. Results

### 3.1. Construction and validation of cell-type-resolved gene regulatory network atlas

To determine whether cell-type-resolved GRNs can be reliably inferred across a large, multi-cohort AD dataset, we constructed a regulatory atlas of the human DLPFC, leveraging the PsychAD Consortium dataset, a single-nucleus transcriptomic resource spanning neurodegenerative and neuropsychiatric disease [21, 31], and assessed its biological coherence and cross-cohort reproducibility (**Figures 1-2; Supplementary Figure 1)**. From this resource, we defined a focused AD phenotyping cohort of 687 individuals, consisting of ∼1.7 million nuclei across 26 transcriptionally defined cell types **(Figure 1; Methods)**.

**Figure 1.**
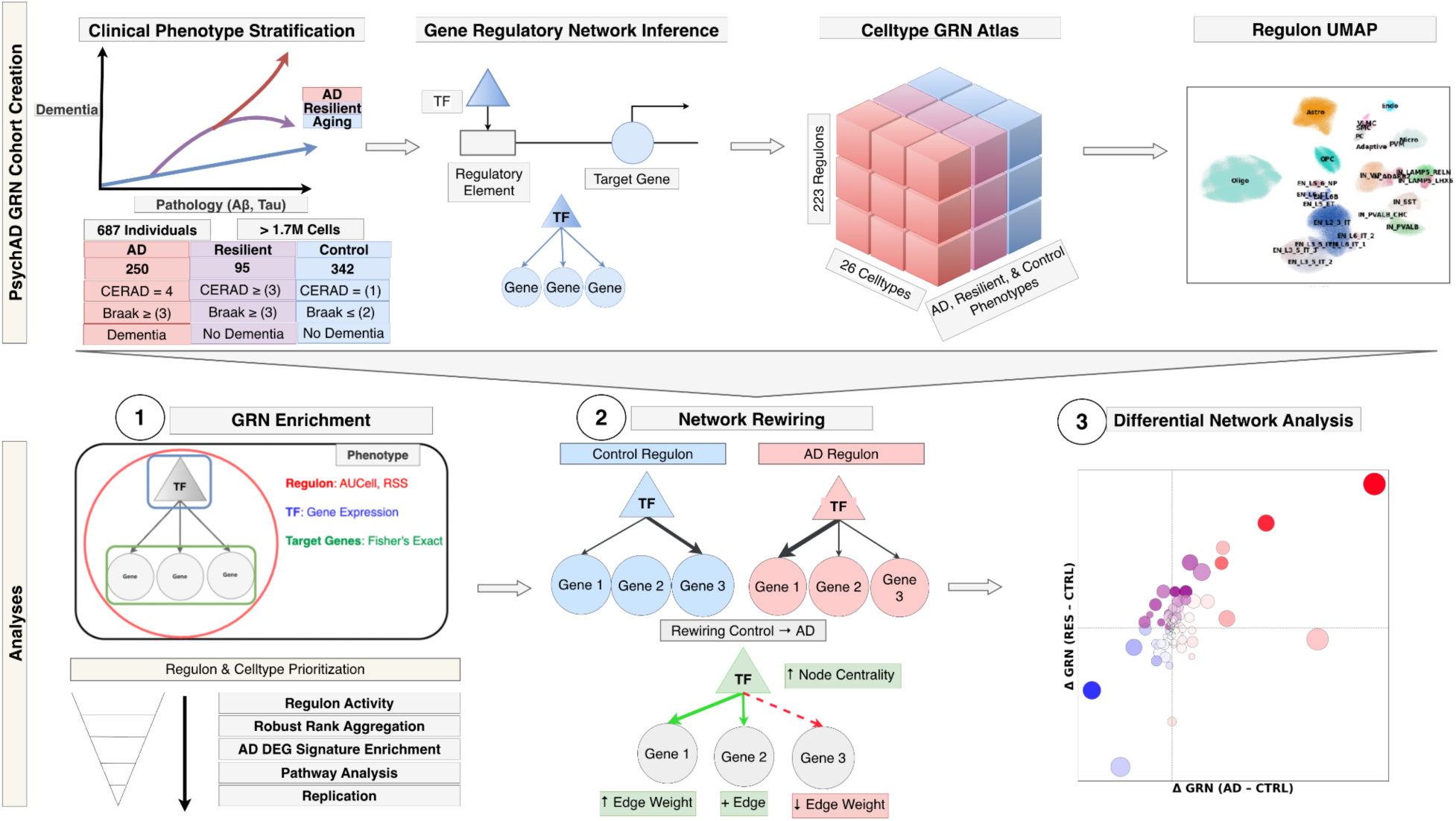
PsychAD GRN Atlas Construction. Single-nucleus RNA-seq profiles from 1.7 million nuclei across 687 individuals from the MSSM (n = 541) and RADC (n = 146) cohorts were annotated into 26 cell types. GRNs were inferred per phenotype (AD, Resilient, and Control) using pySCENIC, and regulon activity (AUCell scores) was aggregated per individual to enable downstream differential and network analyses across phenotypes.

**Figure 2.**
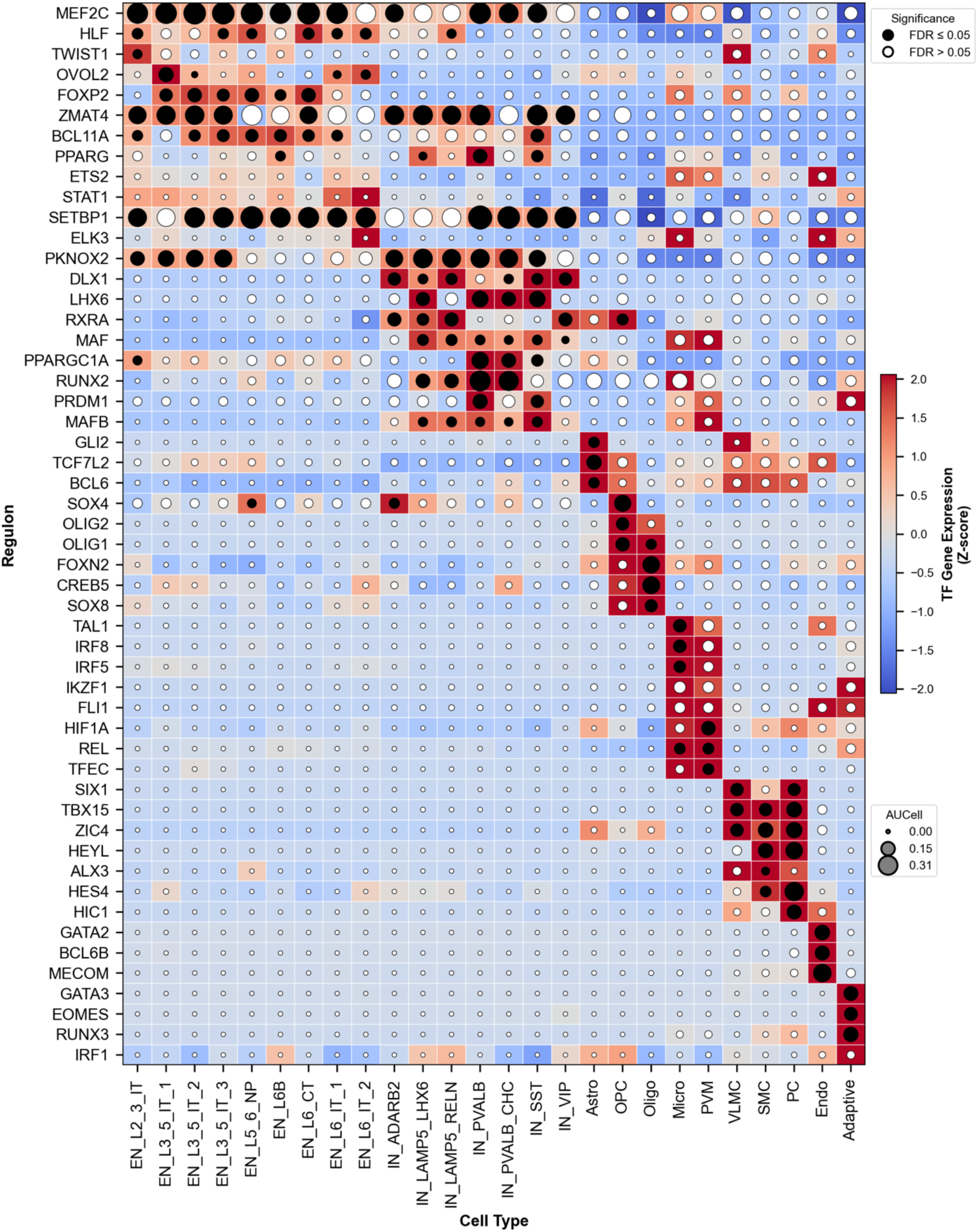
Composite Regulon Prioritization Across Cell Types. Composite heatmap showing the top three regulons per cell type, ranked by Robust Rank Aggregation (RRA). Transcription factor (TF) gene expression Z-scores are shown as square fills, RRA significance as dot fill, and AUCell regulon activity as dot size. Regulons were additionally ranked by regulon specificity score (RSS). This integrated view highlights cell type-specific transcriptional regulators most consistently altered across AD-related analyses.

Using snRNA-seq data from MSSM and RADC cohorts, we constructed a cell-type-resolved GRN atlas **(Figure 1)**. For each phenotype, we inferred TF-target gene regulatory interactions using pySCENIC [25, 26], yielding 223 high-confidence TF regulons. Per-cell regulon activity scores were quantified using AUCell and aggregated to the individual level **(Methods)**, enabling integrative analyses of GRN enrichment and regulatory network rewiring between AD, Resilient, and Control cohorts.

Regulon enrichment analysis supported the biological coherence of the inferred GRN architecture while revealing cell-type-specific patterns relevant to disease **(Figure 2)**. In microglia, IRF8 and TAL1 regulons were strongly enriched, consistent with established myeloid and neuroimmune regulatory programs [32]. BCL6 regulons showed enrichment across glial populations, while vascular and endothelial lineages displayed FLI1 regulon enrichment **(Figure 2)**.

Pathway enrichment analysis [29, 30, 33] of top regulons per cell type revealed distinct lineage-specific functional programs across the DLPFC **(Supplementary Figure 1)**. Neuronal populations were enriched for synaptic signaling, axon guidance, and calcium-dependent processes, reflecting core excitatory and inhibitory functions.

Oligodendrocytes and OPCs showed enrichment for lipid metabolism, myelination, and cholesterol biosynthesis, while astrocytes were associated with oxidative phosphorylation and cytokine signaling. Microglia were enriched for immune activation, interferon response, and phagocytosis, and vascular and perivascular lineages for angiogenesis, extracellular matrix organization, and hypoxia signaling. These patterns confirmed that regulon-defined networks captured canonical, cell type-specific biological pathways, supporting the interpretability and robustness of the PsychAD GRN atlas.

To evaluate reproducibility and generalizability, we compared regulon activity profiles across independent PsychAD cohorts and an external cohort [4] **(Supplementary Figure 2)**. PsychAD MSSM and RADC cohorts showed strong concordance (Pearson r=0.93), and cross-dataset replication demonstrated regulon activity patterns remained highly conserved despite differing cellular taxonomies and sequencing platforms.

### 3.2. Cell-type-specific transcriptional regulators define three key regulatory states across Control, Resilient, and AD groups

We next asked whether organized regulatory configurations distinguish cognitive outcomes across AD, Resilient, and Control groups. To address this, we systematically compared regulon activity, network topology, and regulatory influence **(Figures 3-5)**. We define a regulatory “state” as a coordinated configuration of TF activity characterized by three features: systematic shifts in regulon activity (e.g., AUCell), altered TF-gene connectivity patterns reflecting network rewiring (gain or loss of regulatory edges and edge strength), and changes in regulatory influence on the overall network (PageRank importance within the GRN) **(Methods)**. These states are not mutually exclusive temporal phases or snapshots; rather, they represent regulatory configurations underpinned by distinct molecular interactions across patient groups, with convergence on shared pathways such as NF-κB signaling and cell types, including microglia, astrocytes, and vascular populations.

**Figure 3.**
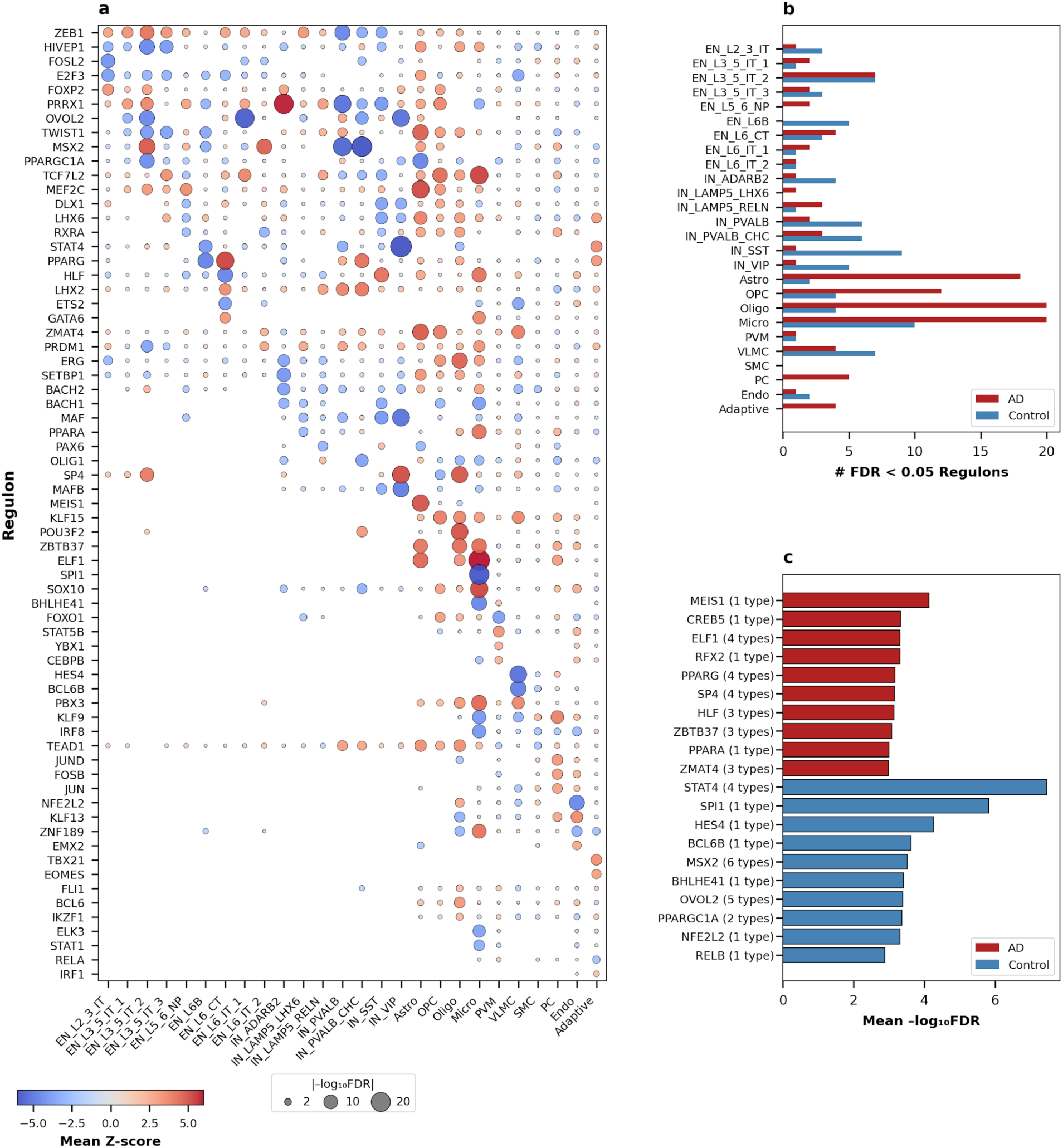
Differential Regulon Activity Across AD Pathology. **(a)** Linear mixed-effects models (Dreamlet) of average differential regulon activity (AUCell Z-score) across MSSM and RADC cohorts for four AD pathology measures (diagnosis, Braak stage, CERAD score, and dementia status; n = 223 regulons tested). Only significant regulons are plotted as individual bubbles, denoting a significant regulon-phenotype association (FDR < 0.05), scaled by effect size and colored by direction of change. **(b)** Number of significant regulons per cell type, split by direction (AD-associated, red; Control-associated, blue). **(c)** Top regulons ranked by mean −log₁₀FDR across AD pathology measures, colored by condition; parenthetical annotations indicate the number of cell types in which each regulon reached significance.

Differential regulon activity analysis across 26 cell types identified 82 regulons significantly associated with AD pathology (FDR < 0.05), with 46 upregulated and 36 downregulated regulons **(Figure 3a)**. Fisher’s exact enrichment testing between regulon target genes and AD differentially expressed genes (derived from [31]) identified 553 significant associations, with 290 AD-enriched and 263 control-enriched **(Figure 4a)**. Network rewiring analysis comparing AD, Resilient, and Control GRNs revealed dramatic reorganization of regulatory architecture **(Figure 5a)**.

**Figure 4.**
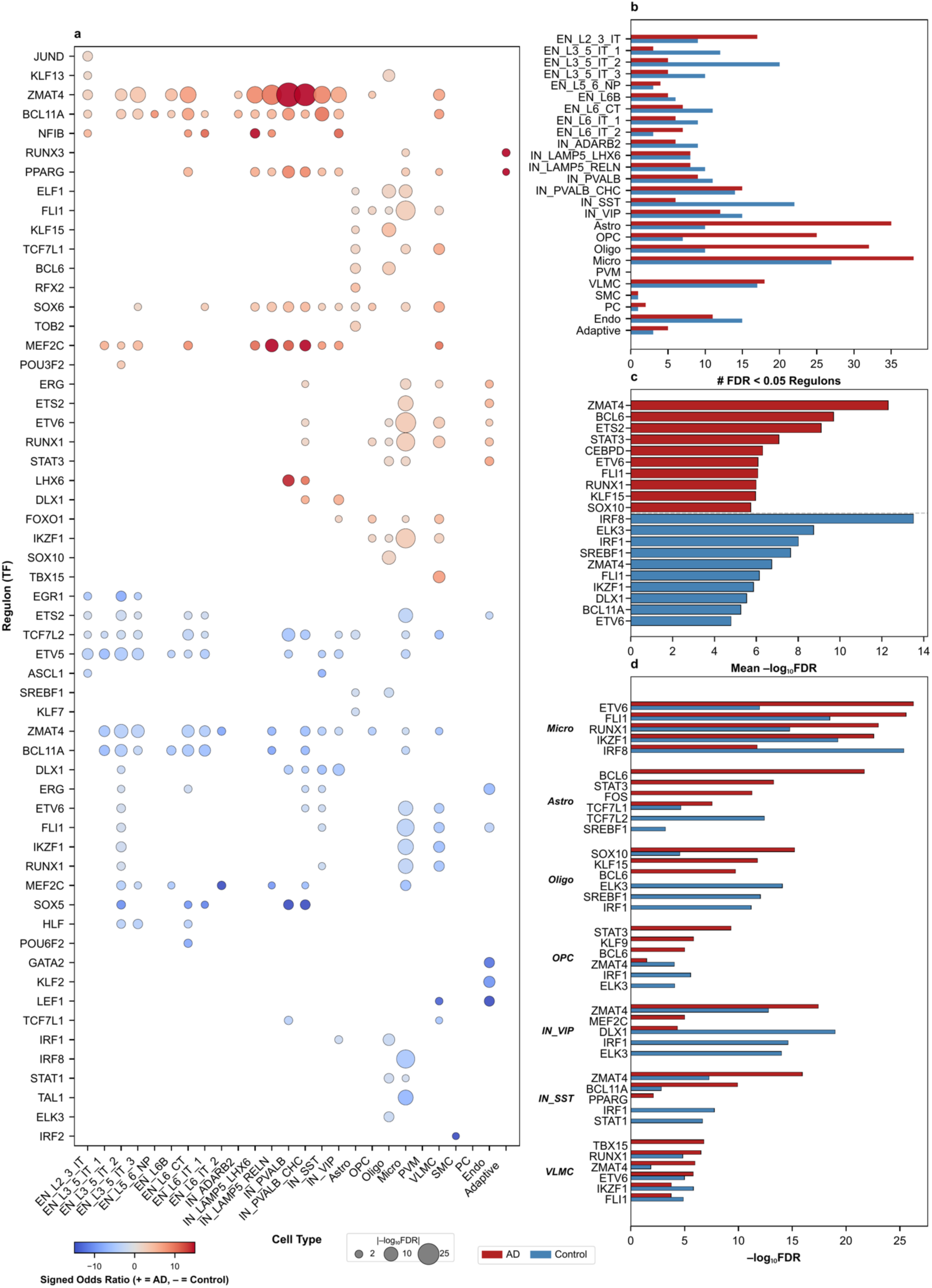
Enrichment of Differentially Expressed Genes in AD and Control Regulons. **(a)** Fisher’s exact test assessing enrichment of AD (red) or control (blue) upregulated differentially expressed genes (DEGs) within regulon target gene sets derived from AD- or control-specific GRNs. Each bubble represents a significant enrichment (FDR < 0.05), scaled by −log_₁₀_FDR. **(b)** Number of significant regulon enrichments per cell type, split by condition. (c) Top regulons ranked by mean−log_₁₀_FDR, with AD-enriched and control-enriched regulons separated by the dashed line. (d) Top three AD- and control-enriched regulons for representative cell types (Micro, Astro, Oligo, OPC, IN_VIP, IN_SST, VLMC), with paired bars showing AD (red) and control (blue) enrichment significance.

#### 3.2.1. State I: Homeostatic Erosion of IRF8/STAT1 Interferon Programs

To identify alteration of homeostatic regulatory programs in AD and the cell types and functional pathways they implicate, we focused on interferon-responsive regulons, particularly IRF8 and STAT1, whose suppression in AD microglia represents the most consistent control-associated regulatory signal in our atlas **(Figures 3-5)**. In State I, we observe systematic suppression of interferon-responsive regulatory programs, particularly in microglia. IRF8, a lineage-determining TF essential for microglial specification from yolk sac erythromyeloid precursors [32], demonstrated a strong disease-associated pattern. We prioritized regulons using Robust Rank Aggregation (RRA; Methods) and identified IRF8 as highly enriched in microglia with the second-highest gene expression z-score (4.31), after TAL1, among all TFs **(Figure 2).** Differential regulon activity revealed broad IRF8 suppression in AD: in microglia (z-score=−3.95, −log₁₀FDR=2.63), with suppression extending to endothelial cells (z-score=−2.64) and smooth muscle cells (z-score=−2.17) **(Figure 3a)**.

Fisher’s exact enrichment testing provided the most striking evidence for IRF8’s role in homeostatic maintenance. IRF8 emerged as the most significantly control-enriched regulon across all TFs and cell types, with microglial enrichment yielding an odds ratio of 5.11 and −log₁₀FDR of 18.27 **(Figure 4a)**. STAT1, a core effector of type I and type II interferon signaling [34], showed coordinated suppression alongside IRF8. Network rewiring analysis revealed that STAT1 underwent the most dramatic loss of regulatory influence, shedding 28 regulatory edges with a −2.63 decrease in network importance **(Figure 5a)**. IRF1 showed parallel contraction, losing 60 edges with a −1.62 decrease in importance. Gene Ontology enrichment of IRF1’s AD-unique targets revealed FDR-significant enrichment for regulation of dendritic cell chemotaxis (FDR = 0.011), negative regulation of immune effector process (FDR = 0.019), ribosomal small subunit assembly (FDR = 0.019), and regulation of adaptive immune response (FDR = 0.019), consistent with IRF1’s established role as an interferon-induced regulator of innate and adaptive immune programs in disease-associated microglia **(Supplementary Figure 3)**. Control-unique targets showed no FDR-significant enrichment, supporting a model in which IRF1 acquires coherent immune-regulatory programs in AD without equivalent acquisition of homeostatic programs in control.

The erosion of homeostatic programs extends beyond microglia to vulnerable neuronal populations. DLX1, essential for interneuron development and specification [35], showed substantial network contraction in AD, losing 35 regulatory edges with −1.22 decrease in importance **(Figure 5a)**. Dreamlet analysis revealed DLX1 downregulation in SST interneurons (z-score=−3.63, −log₁₀FDR=2.19) and VIP interneurons (z-score=−3.34, −log₁₀FDR=1.84), indicating preferential vulnerability of GABAergic populations **(Figure 3a)**. Fisher’s enrichment confirmed control-association across interneuron subtypes: IN_SST (OR=6.06), IN_VIP (OR=5.46), IN_PVALB (OR=4.74), and IN_PVALB_CHC (OR=4.57) **(Figure 4a)**. This broader homeostatic breakdown appears to couple immune activation to circuit-level vulnerability: in PsychAD [31], microglia sit upstream of both vascular remodeling (VLMC increases) and inhibitory interneuron depletion (IN_SST and IN_LAMP5_LHX6), and the inferred directionality suggests that immune escalation is associated with neuronal fragility while simultaneously worsening dementia-linked pathology.

**Figure 5.**
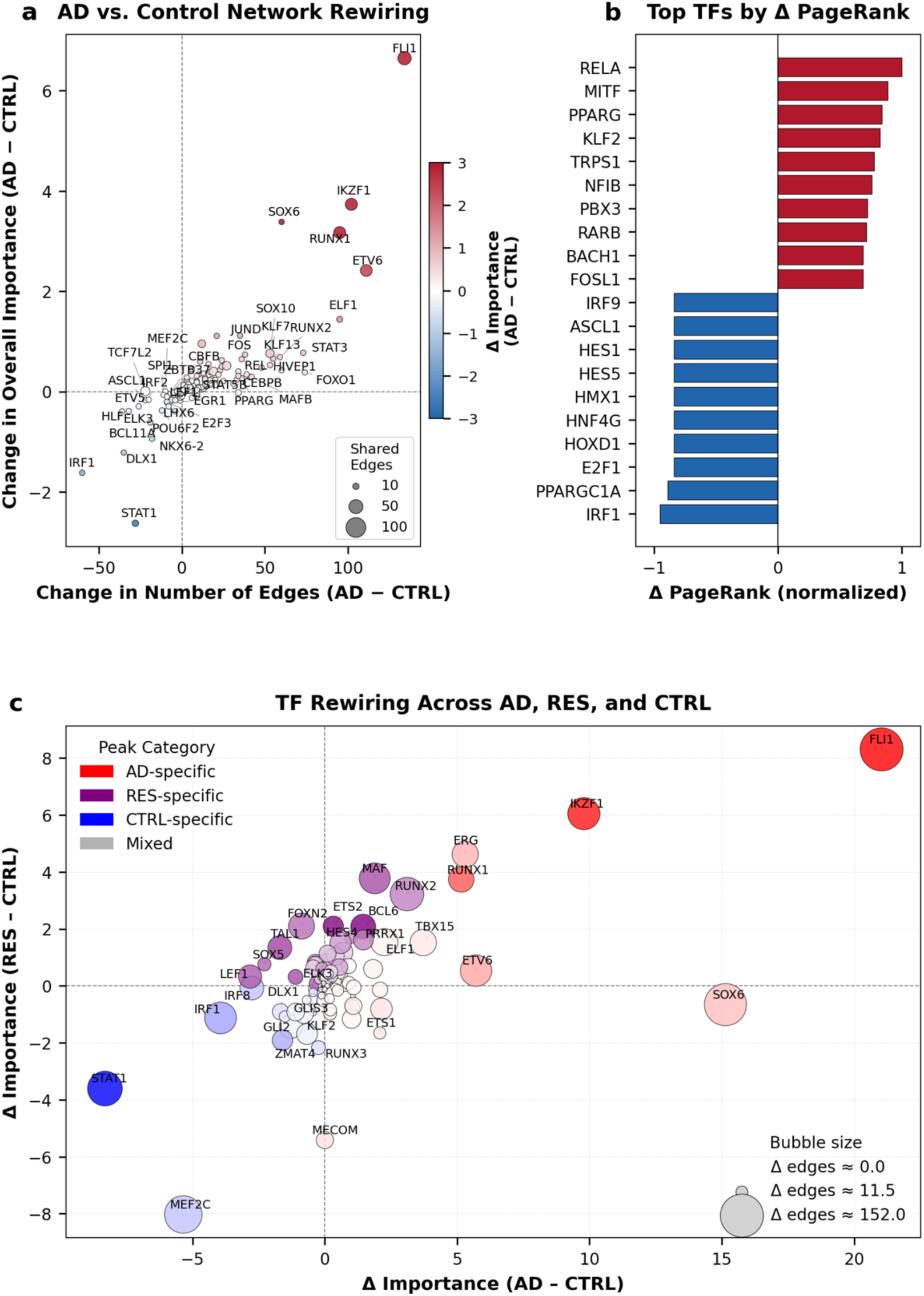
Transcription Factor Network Rewiring and Centrality Shifts in AD. **(a)** Comparison of TF network topology between control and AD GRNs. Scatter plot shows per-TF change in out-degree (Δ edges, x-axis) versus change in aggregate importance of outgoing edges (Δ importance, y-axis) between conditions (AD − CTRL). Point size reflects the number of shared edges between networks, and color indicates direction and magnitude of rewiring. All 174 TFs passing network filters (top 25% of edges; ≥10 target genes) are plotted; the top 20 TFs in each direction (Δ edges) are labeled. **(b)** Top TFs ranked by change in PageRank centrality (AD − CTRL); positive values (red) indicate increased centrality in AD, negative values (blue) indicate decreased centrality. **(c)** Per-TF change in aggregate regulon importance (AD − CTRL, x-axis; RES − CTRL, y-axis), colored by condition of peak importance (AD-specific, red; RES-specific, purple; CTRL-specific, blue). Bubble size reflects magnitude of edge change. TFs in the upper-right quadrant (e.g., FLI1, IKZF1) show increased importance in both AD and Resilient relative to Control; TFs in the lower-left (e.g., STAT1, MEF2C) show decreased importance in both. All 174 TFs passing network filters are plotted; TF labels are limited to the top 10 per peak category (up to 40 total) plus a curated set of key regulators for readability.

#### 3.2.2. State II: Compensatory Resilience Programs Buffer NF-κB via BCL6

We next evaluated whether regulatory programs uniquely engaged in Resilient individuals represent a distinct protective regulatory configuration, or simply an intermediate between Control and AD states. We observed that BCL6 emerged as the strongest resilience-associated regulator, with a distinct network module suggesting active compensation rather than attenuated disease **(Figure 6)**. In State II, a distinct set of TFs is selectively elevated in Resilient individuals. BCL6 demonstrated the strongest Resilient enrichment of any TF, emerging as a molecular switch distinguishing cognitive preservation from decline. Across-group analysis showed BCL6 network importance increased in Resilient relative to Control but decreased in AD relative to Control, consistent with highest aggregate importance in Resilient individuals **(Figure 5c)**. BCL6 showed glial expression across multiple cell types, with highest enrichment in astrocytes (GEX z-score=2.63) alongside oligodendrocyte precursor cells and oligodendrocytes **(Figure 2)**. Dreamlet analysis revealed BCL6 upregulation across ten cell types, with strongest association in oligodendrocytes (z-score=3.20, −log₁₀FDR=1.68) and astrocytes (z-score=1.71) **(Figure 3a)**.

**Figure 6.**
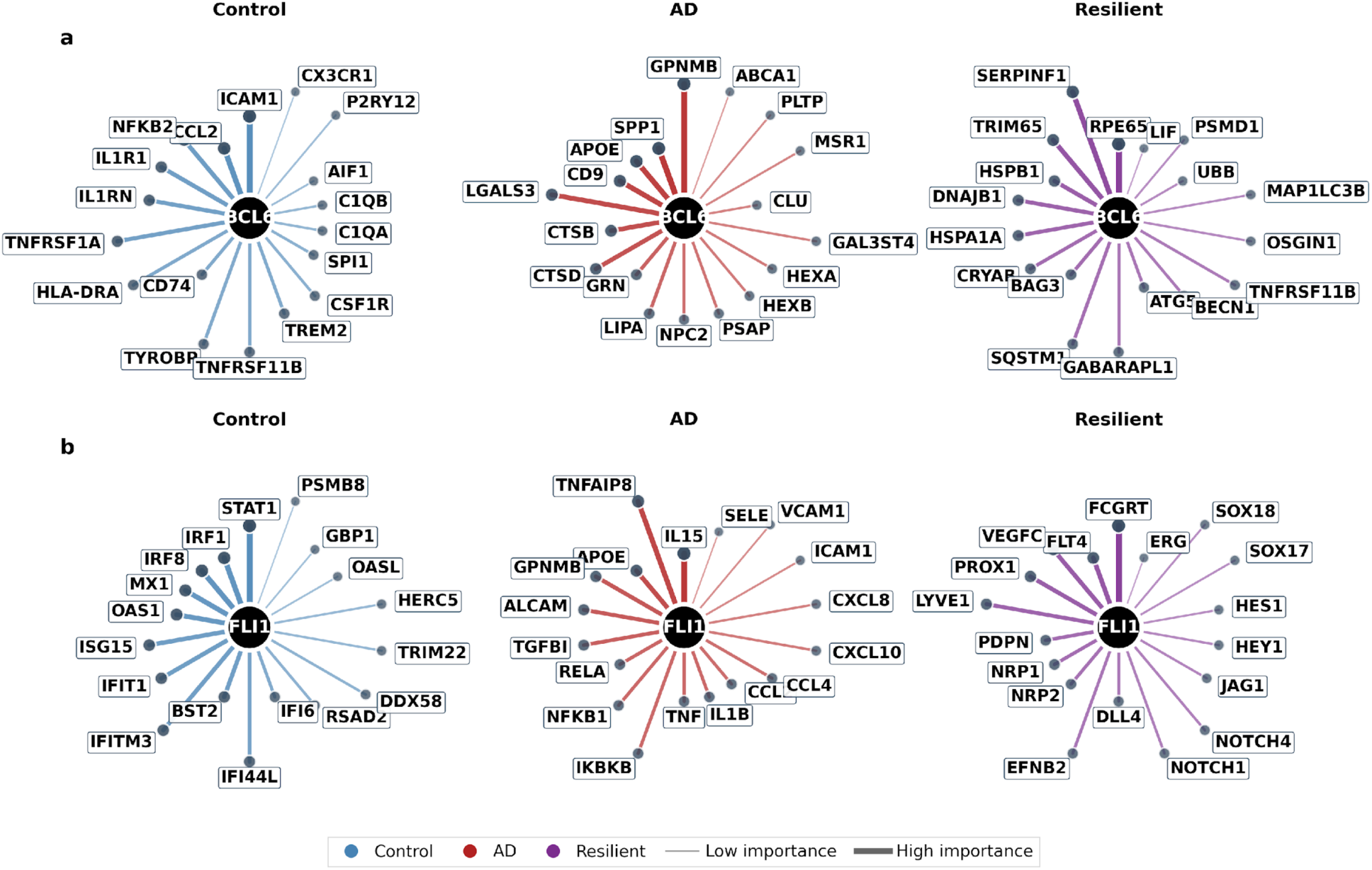
Condition-Specific Regulatory Network Architecture of BCL6 and FLI1. **(a)** BCL6 condition-specific regulatory edges. Circular network layouts display regulatory targets unique to Control (blue, n = 30), AD (red, n = 28), or Resilient (purple, n = 49) networks. Edge width and opacity scale with regulatory importance; node size indicates target gene importance. The 18 highest-importance targets are displayed per panel to preserve label legibility; complete target lists with importance scores are available in the project repository. **(b)** FLI1 condition-specific regulatory edges. Circular network layouts display regulatory targets unique to Control (blue, n = 24), AD (red, n = 85), or Resilient (purple, n = 71) networks. Edge width and opacity scale with regulatory importance; node size indicates target gene importance. The 18 highest-importance targets are displayed per panel to preserve label legibility; complete target lists with importance scores are available in the project repository.

Visualization of condition-specific edges **(Figure 6a)** revealed that BCL6 targets multiple NF-κB pathway components and receptors (NF-κB subunit 2 (NFKB2), interleukin-1 receptor antagonist (IL1R1), tumor necrosis factor receptor 1 (TNFRSF1A)) along with downstream inflammatory genes, consistent with prior evidence that BCL6 and NF-κB occupy overlapping cistromes and exert opposing control over innate immune programs [36]. The positive co-expression of BCL6 with these inflammatory targets, despite BCL6’s canonical repressive function, suggests context-dependent loss of repression or compensatory upregulation in disease states. The BCL6→IL1RN edge was Control-specific and lost in both Resilient and AD, suggesting disruption of an anti-inflammatory regulatory link given IL1RN’s role as the endogenous IL-1 receptor antagonist in glia [37]. Resilient individuals carried the largest BCL6-specific module (49 Resilient-only edges), including TRIM65 [38] and OSGIN1 [39], targets linked to ubiquitin-mediated inflammatory control and oxidative-stress cytoprotection, respectively. By contrast, the BCL6→TNFRSF11B (Osteoprotegerin; OPG) edge was maintained in Resilient but lost in AD; given OPG’s role as a context-dependent regulator of neuroinflammation through the RANK/RANKL/OPG axis [40], this preserved coupling may contribute to anti-inflammatory buffering that distinguishes cognitive resilience from dementia despite comparable neuropathological burden. Relative to Controls, Resilient networks show increased connectivity for BCL6 alongside MAF and RUNX2 **(Supplementary Figure 4b)**, consistent with active compensation through glial-vascular repair programs.

This distinct regulatory architecture was further supported by FLI1 edge analysis, which confirmed selective engagement of adaptive programs alongside active restraint of AD-like expansion, consistent with an alternative regulatory state rather than incomplete disease progression. FLI1 shows fewer Resilient-only than AD-only edges (71 vs. 85) **(Figure 6b)**, but the two sets are qualitatively distinct: Resilient-only targets (e.g., FCGRT, FLT4) implicate enhanced IgG clearance and compensatory lymphangiogenic signaling, whereas AD-only edges concentrate on inflammatory targets. Resilience-unique targets of BCL6 and ETS2 were similarly enriched for blood-brain barrier maintenance and endothelial cell migration, suggesting resilient individuals mount repair responses reminiscent of developmental angiogenesis. Resilient edges are therefore not a subset of AD edges but a distinct configuration, indicating that limiting vascular-immune activation is a defining feature of the resilient state.

#### 3.2.3. State III: Pathogenic Escalation Through FLI1/IKZF1 Network Expansion

Having characterized the Control-associated and Resilient-associated regulatory states, we investigated which regulatory programs undergo the most extensive rewiring in AD and whether they converge on shared inflammatory signaling hubs. Notably, FLI1 and IKZF1 emerged as the most extensively rewired regulators, converging on NF-κB-centered inflammatory programs via expanded network architectures **(Figures 6-7)**. In State III, the AD-associated pattern is characterized by expansion of vascular-immune regulatory networks dominated by ETS family TFs, linked to NF-κB activation and inflammatory remodeling. FLI1 exhibits the most extensive network remodeling among the 174 TFs passing our network filters, gaining 134 regulatory edges in AD relative to Control (67% increase; 6.65-fold importance increase) **(Figure 5a)**. This expansion includes AD-specific regulation of inflammatory and disease-associated genes (IL15, TNFAIP8, APOE, GPNMB [41]) alongside vascular remodeling factors (ALCAM, TGFBI) **(Figure 7a)**. FLI1 showed distributed baseline expression across immune and vascular lineages, concordant with its known role in endothelial development and disease [42, 43], with highest expression in endothelial cells (GEX z-score=3.00), microglia (2.19), and perivascular macrophages (2.14) **(Figure 2).** IKZF1 demonstrates coordinated network expansion with FLI1, gaining 102 regulatory edges (3.73-fold importance increase) in AD relative to Control **(Figure 5a)**. Across the most rewired targets, IKZF1 and FLI1 show substantial convergence (26 of 30 FLI1 targets shared with IKZF1; 87% overlap) **(Figure 7a,b)**, spanning inflammatory mediators (IL15, TNFAIP8), lipid metabolism genes (APOE), and vascular remodeling programs (TGFBI). PageRank centrality analysis revealed that RELA (NF-κB p65) shows the greatest gain among the TFs that pass filters in network centrality in AD **(Figure 5b)**, positioning NF-κB signaling at the center of rewired AD networks. The opposition between State II BCL6-mediated NF-κB suppression and State III FLI1/RELA-mediated NF-κB activation may represent a molecular switch distinguishing cognitive resilience from dementia. The contrast in edge dynamics is stark: State I regulators IRF8, IRF1, and STAT1 collectively lost substantial numbers of edges (IRF1: 60, STAT1: 28), while State III regulators FLI1 and IKZF1 gained approximately 236 edges, a differential of over 300 regulatory connections reflecting fundamental reorganization of the brain’s transcriptional landscape across AD-associated regulatory states.

**Figure 7.**
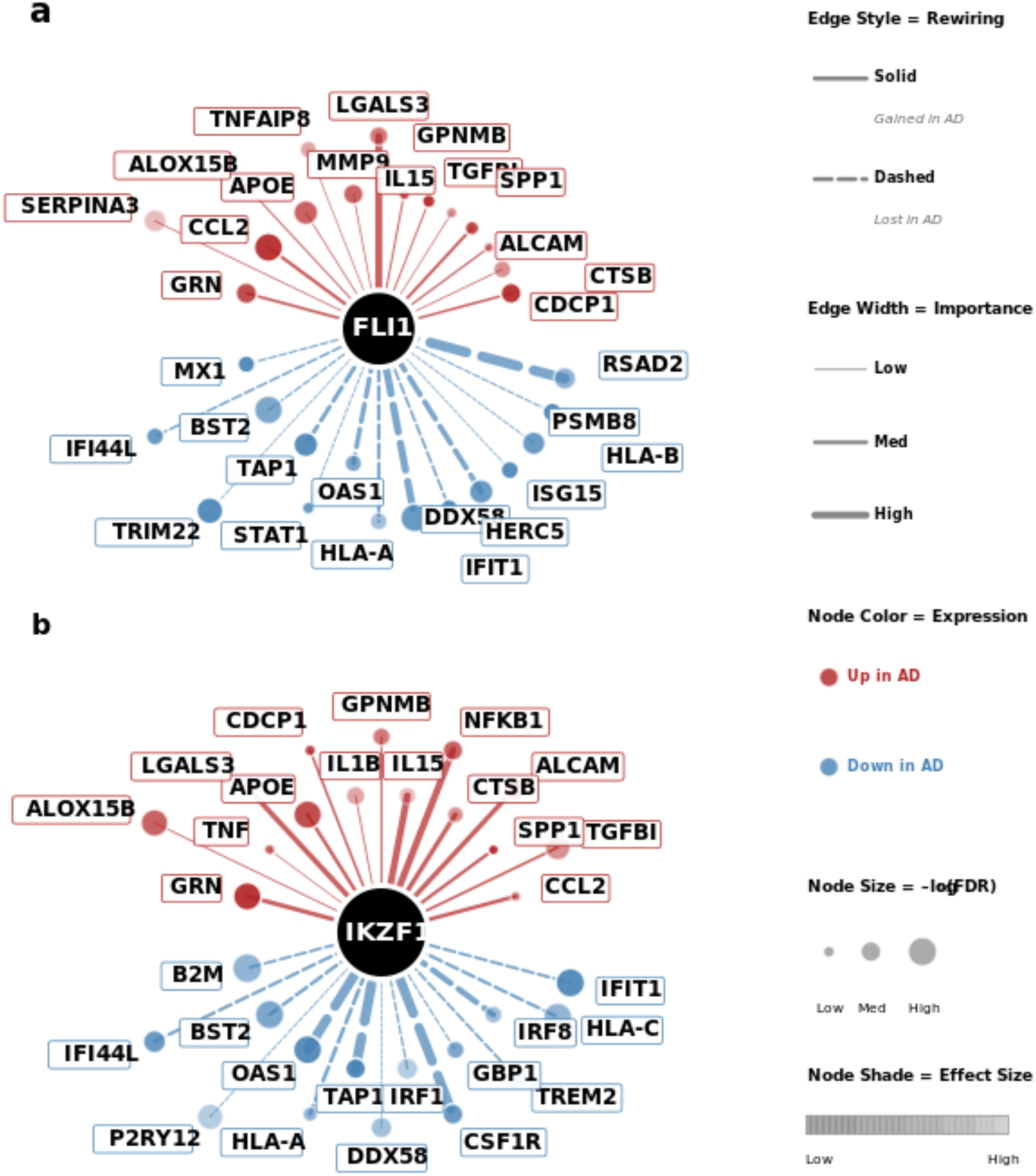
Network Rewiring of FLI1 and IKZF1 in AD. **(a)** FLI1 regulatory network rewiring comparing AD versus Control. Circular layout shows 30 rewired targets (stable edges excluded). Top hemisphere: targets gained in AD (red nodes, solid edges, n = 15); Bottom hemisphere: targets lost in AD (blue nodes, dashed edges, n = 15). Edge width scales with regulatory strength; node size with differential expression significance (−log₁₀FDR); node intensity with effect size magnitude. **(b)** IKZF1 regulatory network rewiring comparing AD versus Control. Circular layout shows 30 rewired targets. Top hemisphere: targets gained in AD (red nodes, solid edges, n = 15); bottom hemisphere: targets lost in AD (blue nodes, dashed edges, n = 15). Edge width scales with regulatory strength; node size with expression significance; node intensity with effect size.

Integration of differential activity, enrichment, and network analyses converged on a consistent three-state framework. Control-associated regulators IRF8, STAT1, and IRF1 uniformly showed downregulation in AD (mean dreamlet z-score=−2.85), strong control-enrichment (mean Fisher’s OR=3.17), and network contraction (edge loss=49). Resilient-associated regulators BCL6, TAL1, and ETS2 showed cell-type-specific patterns with highest importance in resiliency, moderate enrichment in both conditions, and distinct network metrics reflecting unique regulatory engagement. AD-associated regulators FLI1 and IKZF1 showed upregulation trends, strong AD-enrichment (mean OR=2.97), and massive network expansion (mean edge gain=118). Microglia harbored the most disease-associated regulons (30 total), alongside oligodendrocytes (24), astrocytes (20), and OPCs (16), establishing glial populations as the primary locus of transcriptional dysregulation in AD.

## 4. Discussion

This study presents a comprehensive cell-type-resolved GRN atlas of AD, comparing Control, Resilient, and AD dementia groups. By systematically comparing regulatory network architecture across these groups in 687 individuals and 1.7 million nuclei [21, 31], we identify a consistent organizing principle: the balance between homeostatic programs, resilient-associated compensation, and inflammatory activation aligns with cognitive outcomes in the face of neuropathology. Our three-state framework provides a conceptual model for understanding how transcriptional regulatory dynamics distinguish cognitive outcomes despite similar pathological burden. While these states are identified through cross-sectional comparisons, they represent distinct regulatory configurations associated with divergent cognitive outcomes. Individuals with comparable neuropathological burden engage fundamentally different regulatory architectures, with resilience representing an independent regulatory state rather than an intermediate stage. BCL6 emerges as a molecular switch of protective and pathogenic responses, whose regulatory activity distinguishes resilient from pathogenic states.

The atlas identified TFs whose activity patterns recapitulate canonical developmental hierarchies — including cortical interneuron specification [35], neuronal FOXP2 expression [44], and oligodendrocyte OLIG function [45] — validating the biological coherence of inferred networks. Many regulators exhibiting dramatic disease-associated rewiring are lineage-determining factors establishing cellular identity during development. IRF8 and PU.1 direct microglial specification from yolk sac erythromyeloid precursors [32], while FLI1 governs hemangioblast specification and endothelial fate decisions [42, 43]. Notably, our IRF8-centered microglial signature aligns with recent single-nucleus studies demonstrating IRF8-driven reactive microglia phenotypes in human AD that differ markedly from the disease-associated microglia (DAM) signature characterized in mouse models. Zhou et al. [46] reported that human AD microglia exhibit an IRF8-driven reactive signature distinct from the TREM2-dependent DAM response, consistent with our finding that IRF8 activity distinguishes Control from AD states.

NF-κB regulatory balance emerges as a unifying mechanism across our three-state framework. Control-associated interferon regulators IRF1 and STAT1 cooperate with NF-κB in tightly controlled immune responses under homeostatic conditions [47–49]; reduced activity of these regulators in AD may destabilize this regulatory crosstalk. RELA, a core NF-κB subunit, showed the greatest gain in network centrality in AD **(Figure 5b)**, suggesting reinforcement of an NF-κB-centered regulatory architecture, with increased coupling of inflammatory signaling to rewired transcriptional programs. The condition-specific edges **(Figure 6a,b)** highlight opposing BCL6- and FLI1-centered programs that differentiate resilience from dementia. BCL6 shows the largest Resilient-specific module and retains regulation of anti-inflammatory pathway genes such as TNFRSF11B — notably, TNFRSF11B encodes OPG, a decoy receptor with context-dependent roles in neuroinflammation [40], whereas FLI1 displays a larger AD-specific expansion, supporting vascular-immune inflammatory remodeling.

IRF1 AD-unique targets showed FDR-significant enrichment for innate and adaptive immune-regulatory programs **(Supplementary Figure 3)**, while no IRF1 control-unique target term reached FDR significance, consistent with a transition of IRF1 from a constrained homeostatic regulon in control toward an expanded immune-regulatory program in AD. The reduction of IRF8 and IRF1 network centrality together with increased FLI1 expansion reflects a contrast between Control-associated homeostatic programs and AD-associated inflammatory activation.

This finding resonates with emerging evidence from Udeochu et al. [50] demonstrating that innate immune activation through cGAS-STING signaling diminishes cognitive resilience by decreasing neuronal MEF2C expression through type I interferon signaling. Their work supports a model in which type I interferon dysregulation contributes to the erosion of cognitive resilience, thereby enabling progression to symptomatic dementia; this is consistent with our model that attenuation of the IRF8/STAT1/IRF1 regulatory axis represents a loss of homeostatic buffering capacity, which may precede the downstream pathogenic escalation observed in State III.

Resilient-associated regulatory programs revealed coordinated glial-vascular responses distinguishing cognitively preserved individuals. BCL6 exhibited the strongest network importance enrichment in Resilient individuals, alongside MAF, RUNX2, ETS2, and ELK3, with enrichment for inflammatory regulation, microglial activation, and vascular remodeling. Resilient-enriched BCL6 activation in glia may enable adaptation without full commitment to reactive inflammatory states. BCL6 is known to suppress NF-κB signaling and drive neurogenic fate transitions by inhibiting self-renewal pathways [36, 51]. Our identification of BCL6-mediated glial buffering provides a complementary mechanism operating in non-neuronal populations.

The enrichment of BCL6 and ETS2 resilience-unique targets for vascular maintenance pathways, combined with the constrained FLI1 network architecture in resilience, supports a model in which limiting vascular-immune activation preserves homeostasis. Together, these findings position the BCL6-FLI1 regulatory axis as a molecular switch determining whether glial responses resolve toward homeostasis (maintained BCL6 activity) or escalate toward chronic inflammation (FLI1/RELA expansion).

Our findings highlight potential therapeutic targets positioned along the three-state framework. Sustaining State I IRF8/STAT1 homeostatic programs may preserve microglial surveillance. IRF8’s emergence as the most significantly control-enriched regulon (microglial OR=5.11, −log₁₀FDR=18.27) identifies it as a critical node whose maintenance may counter AD-associated regulatory states. BCL6 represents a molecular switch whose sustained activation could prolong the resilient state by maintaining NF-κB suppression [36]. Constraining State III FLI1-driven vascular-immune activation could stabilize blood-brain barrier integrity and limit NF-κB-mediated inflammation. Most critically, the BCL6 elevation that distinguishes Resilient individuals suggests interventions extending State II adaptive responses during early pathology might promote cognitive preservation.

Several limitations warrant consideration. GRN inference from single-cell transcriptomics captures correlational rather than causal relationships; perturbation experiments in model systems will be essential for validation. Critically, we cannot determine from cross-sectional data whether the regulatory shifts we observe drive disease progression, represent compensatory responses to pathology, or are downstream consequences of other pathogenic processes. For example, the erosion of IRF8/STAT1 programs in AD could reflect a causal loss of homeostatic surveillance that permits pathogenic escalation, or it could be a secondary consequence of upstream events such as chronic Aβ exposure or metabolic stress. Similarly, BCL6 activation in resilient individuals may actively protect cognition or may be a marker of a broader protective biology not fully captured by our GRN framework. Resolving this directionality and validating BCL6 as a functional mediator of resilience, rather than merely a correlate, represents the highest-priority future experiment for solidifying the three-state framework. Our cross-sectional design cannot establish temporal ordering of transcriptional changes; future work leveraging continuous pathology measures and longitudinal cohorts may clarify whether the three states represent sequential transitions or co-occurring regulatory programs whose balance shifts with disease.

Additional methodological constraints merit explicit acknowledgment. Our analyses rely on post-mortem tissue, which provides terminal snapshots influenced by agonal state, perimortem medication exposure, and post-mortem interval, and cannot capture the dynamic trajectory of regulatory changes during disease progression in a living brain. The GRN inference approach combines co-expression patterns with TF motif priors and therefore captures statistical associations rather than direct physical TF-target interactions. It cannot distinguish direct from indirect regulatory relationships, nor can it resolve whether co-expression reflects shared upstream regulation rather than a direct TF-target relationship. As an immediate next step, integration with existing AD single-nucleus ATAC-seq (snATAC-seq) or multiome data will be essential to validate our highest-priority network edges through direct chromatin accessibility evidence.

Furthermore, our cohorts are drawn predominantly from individuals of European ancestry, and findings from the DLPFC may not generalize to other brain regions or demographic populations. Whether the three-state framework generalizes to non-European populations or other cortical and subcortical regions affected in AD remains to be tested. Despite these constraints, the convergence of multiple analytical approaches (e.g. differential activity, Fisher’s enrichment, and network topology) across two independent cohorts (MSSM and RADC) with distinct ascertainment and technical characteristics, together with replication in an external cohort [4], supports the robustness and generalizability of the core three-state framework.

In summary, this atlas reveals that cognitive resilience in the face of AD neuropathology is defined not by attenuated pathology or intermediate disease progression, but by active modulation of immune and vascular-associated regulatory programs. Resilient individuals engage a distinct glial-vascular regulatory configuration, anchored by BCL6-mediated suppression of NF-κB signaling within a constrained FLI1 network architecture that preserves anti-inflammatory coupling and blood-brain barrier maintenance even as homeostatic interferon surveillance erodes. These findings reframe dementia as a failure of cell-type-specific regulatory networks to sustain homeostatic and resilience-associated programs at the neurovascular niche. The balance between BCL6 suppression and FLI1/RELA expansion in glial and endothelial populations thus emerges as a candidate regulatory mechanism underlying divergent cognitive outcomes in AD.

## Author information

### PsychAD Consortium Authors

Aram Hong (1, 2, 3, 4); Athan Z. Li (15, 17); Biao Zeng (1, 2, 3, 4); Chenfeng He (14, 17); Chirag Gupta (14, 17); Christian Dillard (1, 2, 3, 4); Christian Porras (1, 2, 3, 4); Clara Casey (1, 2, 3, 4); Colleen A. McClung (23); Collin Spencer (1, 2, 3, 4); Daifeng Wang (14, 15, 17); David A. Bennett (9, 10); David Burstein (1, 2, 3, 4, 6, 7); Deepika Mathur (1, 2, 3, 4, 6, 7); Donghoon Lee (1, 2, 3, 4); Fotios Tsetsos (1, 2, 3, 4, 6); Gabriel E. Hoffman (1, 2, 3, 4, 6, 7); Genadi Ryan (18, 22); Georgios Voloudakis (1, 2, 3, 4, 6, 7, 12); Hui Yang (1, 2, 3, 4); Jaroslav Bendl (1, 2, 3, 4); Jerome J. Choi (16, 17); John F. Fullard (1, 2, 3, 4); Kalpana H. Arachchilage (14, 17); Karen Therrien (1, 2, 3, 4); Kiran Girdhar (1, 2, 3, 4); Lars J. Jensen (5); Lisa L. Barnes (9, 10); Logan C. Dumitrescu (24, 25); Lyra Sheu (1, 2, 3, 4); Madeline R. Scott (23); Marcela Alvia (1, 2, 3, 4); Marios Anyfantakis (1, 2, 3, 4); Maxim Signaevsky (2, 3); Mikaela Koutrouli (1, 2, 3, 4, 5); Milos Pjanic (1, 2, 3, 4); Monika Ahirwar (18, 22); Nicolas Y. Masse (1, 2, 3, 4); Noah Cohen Kalafut (15, 17); Panos Roussos (1, 2, 3, 4, 6, 7); Pavan K. Auluck (8); Pavel Katsel (3); Pengfei Dong (1, 2, 3, 4); Pramod B. Chandrashekar (14, 17); Prashant N.M. (1, 2, 3, 4); Rachel Bercovitch (1, 2, 3, 4); Roman Kosoy (1, 2, 3, 4); Sanan Venkatesh (1, 2, 3, 4, 6); Saniya Khullar (14, 17); Sarah R. Murphy (1, 2, 3, 4); Sayali A. Alatkar (15, 17); Seon Kinrot (1, 2, 3, 4); Stathis Argyriou (1, 2, 3, 4); Stefano Marenco (8); Steven Finkbeiner (18, 19, 20, 21, 22); Steven P. Kleopoulos (1, 2, 3, 4); Tereza Clarence (1, 2, 3, 4); Timothy J. Hohman (24, 25); Ting Jin (14, 17); Vahram Haroutunian (2, 3, 7, 13); Vivek G. Ramaswamy (18, 22); Xiang Huang (17); Xinyi Wang (1, 2, 3, 4); Zhenyi Wu (1, 2, 3, 4); Zhiping Shao (1, 2, 3, 4)

### PsychAD Consortium Affiliations

1: Center for Disease Neurogenomics, Icahn School of Medicine at Mount Sinai, New York, NY, USA.

2: Friedman Brain Institute, Icahn School of Medicine at Mount Sinai, New York, NY, USA.

3: Department of Psychiatry, Icahn School of Medicine at Mount Sinai, New York, NY, USA.

4: Department of Genetics and Genomic Sciences, Icahn School of Medicine at Mount Sinai, New York, NY, USA.

5: Novo Nordisk Foundation Center for Protein Research, Faculty of Health and Medical Sciences, University of Copenhagen, Copenhagen, Denmark.

6: Center for Precision Medicine and Translational Therapeutics, James J. Peters VA Medical Center, Bronx, NY, USA.

7: Mental Illness Research, Education and Clinical Center VISN2, James J. Peters VA Medical Center, Bronx, NY, USA.

8: Human Brain Collection Core, National Institute of Mental Health-Intramural Research Program, Bethesda, MD, USA.

9: Rush Alzheimer’s Disease Center, Rush University Medical Center, Chicago, Illinois, USA.

10: Department of Neurological Sciences, Rush University Medical Center, Chicago, Illinois, USA.

11: PsychAD Consortium

12: Department of Artificial Intelligence and Human Health, Icahn School of Medicine at Mount Sinai, New York, NY, USA.

13: Department of Neuroscience, Icahn School of Medicine at Mount Sinai, New York, NY, USA.

14: Department of Biostatistics and Medical Informatics, University of Wisconsin-Madison, Madison, WI, USA.

15: Department of Computer Sciences, University of Wisconsin-Madison, Madison, WI, USA.

16: Department of Population Health Sciences, University of Wisconsin-Madison, Madison, WI, USA.

17: Waisman Center, University of Wisconsin-Madison, Madison, WI, USA.

18: Center for Systems and Therapeutics, Gladstone Institutes, San Francisco, CA, USA.

19: Department of Neurology, University of California San Francisco, San Francisco, CA, USA.

20: Department of Physiology, University of California San Francisco, San Francisco, CA, USA.

21: Neuroscience and Biomedical Sciences Graduate Programs, University of California San Francisco, San Francisco, CA, USA.

22: Taube/Koret Center for Neurodegenerative Disease Research, Gladstone Institutes, San Francisco, CA, USA.

23: Department of Psychiatry, University of Pittsburgh School of Medicine, Pittsburgh, PA, USA.

24: Vanderbilt Genetics Institute, Vanderbilt University Medical Center, Nashville, TN, USA.

25: Vanderbilt Memory & Alzheimer’s Center, Vanderbilt University Medical Center, Nashville, TN, USA.

## Contributions

Conceptualization: Collin Spencer (CS), Donghoon Lee (DL), John F. Fullard (JFF)

Panos Roussos (PR)

Methodology: CS, DL, Gabriel E. Hoffman (GEH), JFF, PR

Software: CS, GEH

Validation: CS

Formal analysis: CS, GEH

Investigation: CS, JFF

Resources: DL, PR

Data Curation: CS, DL, Prashant N.M. (PNM), Jaroslav Bendl (JB)

Writing: CS, DL, PR

Visualization: CS

Supervision: DL, JB, JFF, PR

Project administration: DL, PR

Funding acquisition: DL, PR

All authors read and approved the final draft of the paper.

## Data Availability

Raw single-nucleus RNA-seq data are available via the AD Knowledge Portal (https://doi.org/10.7303/9618136). The AD Knowledge Portal is a platform for accessing data, analyses, and tools generated by the Accelerating Medicines Partnership (AMP-AD) Target Discovery Program and other National Institute on Aging (NIA)-supported programs to enable open-science practices and accelerate translational learning. The data, analyses and tools are shared early in the research cycle without a publication embargo on secondary use. Data is available for general research use according to the following requirements for data access and data attribution (https://adknowledgeportal.synapse.org/Data Access). Derived GRN results, including regulon definitions, AUCell enrichment matrices, and network edge lists, are available alongside analysis code at https://github.com/DiseaseNeuroGenomics/PsychAD_GRN.

## Acknowledgments

We would like to express our deep gratitude to the patients and their families who generously donated the invaluable biological material essential for the success of this study. We are profoundly indebted to their participation and commitment to advancing scientific knowledge and improving human health. The results published here are in whole or in part based on data obtained from the AD Knowledge Portal.

The results published here are in whole or in part based on data obtained from the The AD Knowledge Portal (https://doi.org/10.7303/9618136). Study data were generated from the postmortem brain tissue provided by the Mount Sinai Brain Bank (MSBB), Rush Alzheimer’s Disease Center (RUSH; funding: P30AG10161, P30AG72975, R01AG15819, R01AG17917. U01AG46152, and U01AG61356), and NIMH-IRP Human Brain Collection Core (HBCC, project # ZIC MH002903). The MSBB specimens were provided through the NIH NeuroBioBank and supported by NIMH-75N95019C00049. Data collection was supported through funding by NIA grants R01AG067025 and was provided through the work of PIs Panos Roussos (Icahn School of Medicine at Mount Sinai), Vahram Haroutunian (Icahn School of Medicine at Mount Sinai), Steven Finkbeiner (Gladstone Institutes), Daifeng Wang (University of Wisconsin-Madison).

Study data from The Religious Orders Study and Memory and Aging Project (ROSMAP) Study (https://doi.org/10.7303/9618239) were provided by the Rush Alzheimer’s Disease Center, Rush University Medical Center, Chicago. Data collection was supported through funding by NIA grants P30AG10161 (ROS), R01AG15819 (ROSMAP; genomics and RNAseq), R01AG17917 (MAP), R01AG30146, R01AG36042 (5hC methylation, ATACseq), RC2AG036547 (H3K9Ac), R01AG36836 (RNAseq), R01AG48015 (monocyte RNAseq) RF1AG57473 (single nucleus RNAseq), U01AG32984 (genomic and whole exome sequencing), U01AG46152 (ROSMAP AMP-AD, targeted proteomics), U01AG46161(TMT proteomics), U01AG61356 (whole genome sequencing, targeted proteomics, ROSMAP AMP-AD), P30AG072975, the Illinois Department of Public Health (ROSMAP), and the Translational Genomics Research Institute (genomic). snRNA-seq data generation was funded by NIH grants U01AG061356 (De Jager/Bennett), RF1AG057473 (De Jager/Bennett), and U01AG046152 (De Jager/Bennett) as part of the AMP-AD consortium, as well as NIH grants R01AG066831 (Menon) and U01AG072572 (De Jager/St George-Hyslop). Additional phenotypic data can be requested at www.radc.rush.edu.

## Consent Statement

All brain specimens were obtained through informed consent via brain donation programs. This study was approved by the Institutional Review Boards of the Icahn School of Medicine at Mount Sinai, the National Institute of Mental Health, and Rush University Medical Center, and was performed in accordance with the ethical standards of the 1964 Declaration of Helsinki and its later amendments. To address the heterogeneity of AD and at-risk populations, the PsychAD cohort was sampled to span diverse genetic ancestries, sexes, ages, and neuropathological stages.

## Materials & Correspondence

Correspondence to Collin Spencer, Donghoon Lee, and Panos Roussos. Mailing address: 1470 Madison Avenue, Hess Center for Science and Medicine, Floor 9, Room 107, New York, NY 10029, USA.

## Supplementary Figures

**Supplementary Figure 1.**
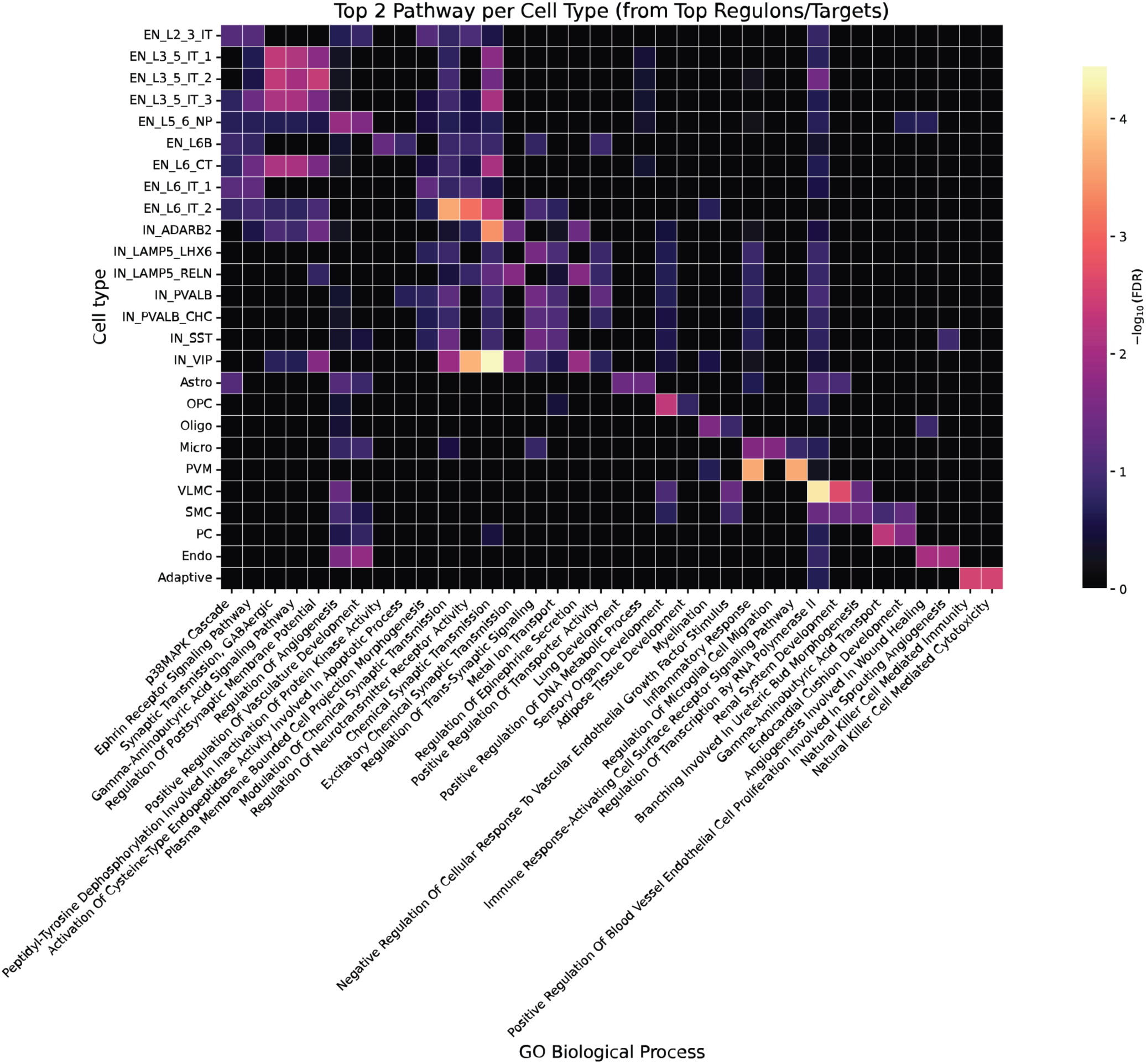
Regulon target gene pathway enrichment. Gene Ontology Biological Process enrichment for each cell type using target genes from the top 3 highest-priority regulons per cell type; top two enriched pathways shown per cell type.

**Supplementary Figure 2.**
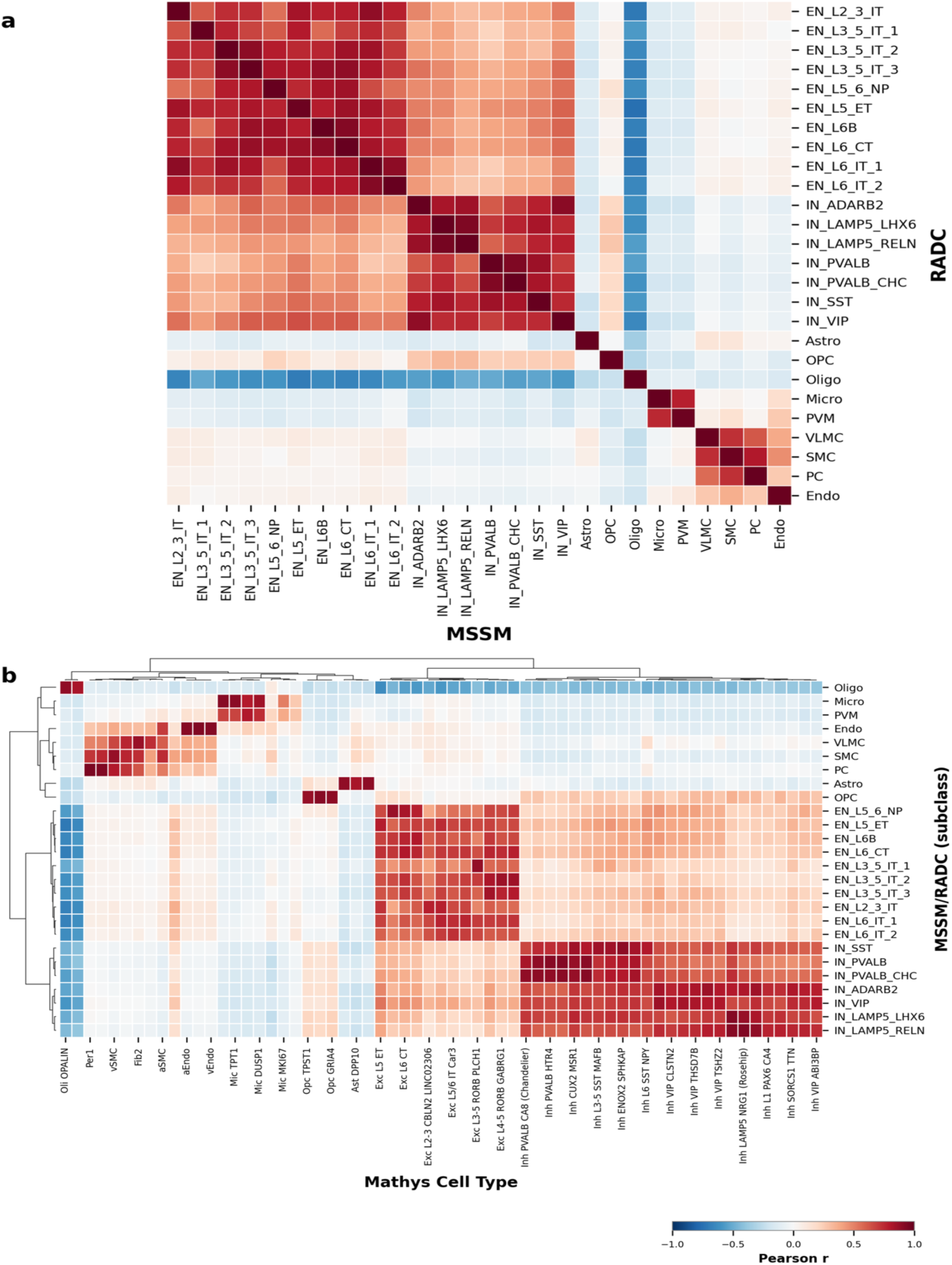
Cross-dataset regulon activity alignment. **(a)** Pearson correlation matrix comparing cell type-level regulon activity (AUCell) between MSSM and RADC cohorts. **(b)** Hierarchically clustered heatmap of Z-scored regulon activity across matched MSSM/RADC and external reference (Mathys) cell-type signatures, illustrating broad cross-dataset alignment.

**Supplementary Figure 3.**
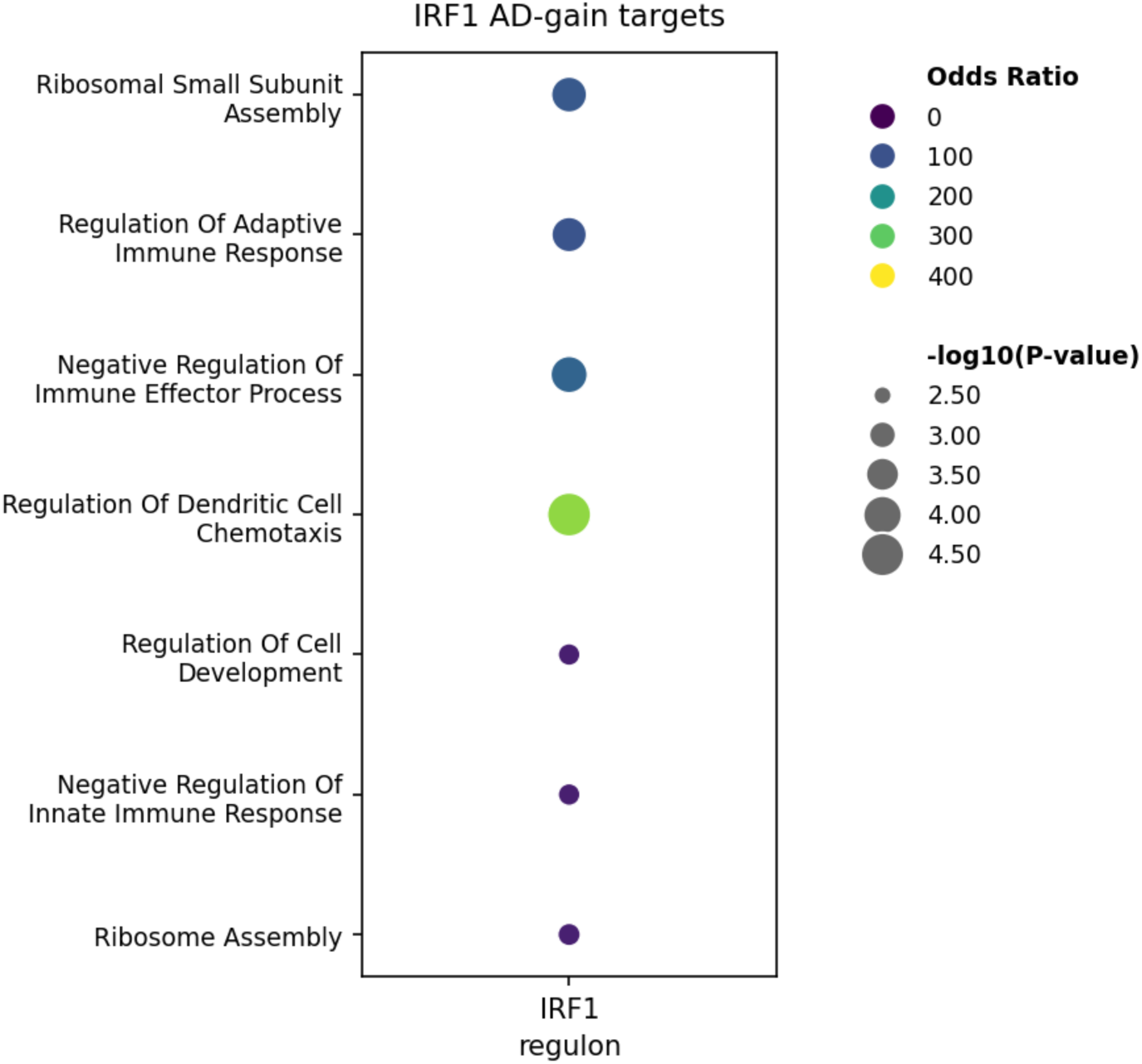
IRF1 AD-gain regulon target gene GO enrichment. Gene Ontology Biological Process enrichment of IRF1 target genes unique to the AD GRN. Dot size encodes −log₁₀(P-value); color encodes odds ratio. Four terms reach FDR < 0.05; the corresponding control-gain analysis yielded no FDR-significant terms.

**Supplementary Figure 4.**
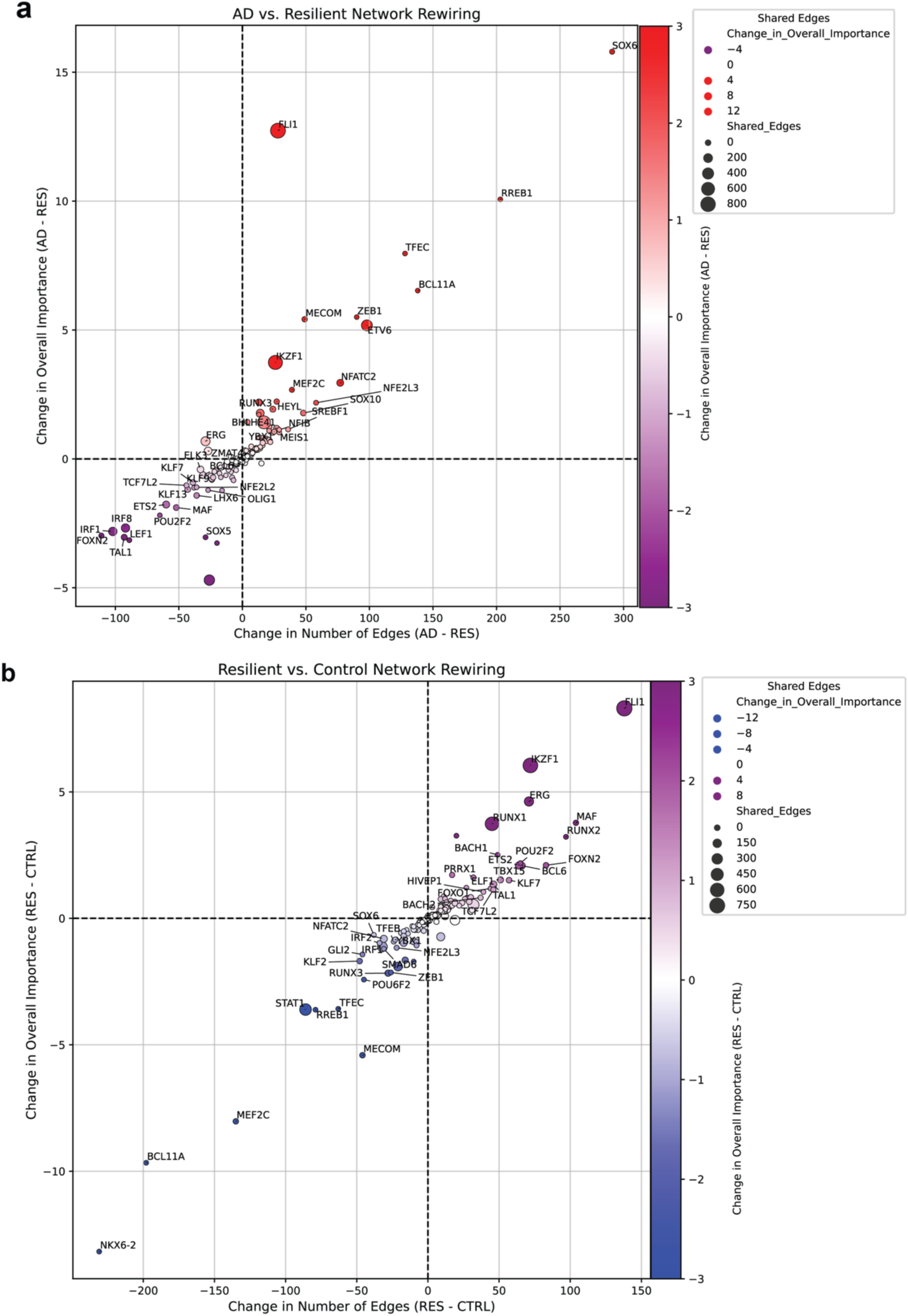
Network rewiring across resilient and disease states. **(a)** AD vs. Resilient network rewiring. Per-TF change in out-degree (Δ edges, x-axis) versus change in aggregate importance (Δ importance, y-axis). Point size = shared edges; color = magnitude/direction. **(b)** Resilient vs. Control network rewiring (same encoding).

